# Fractal and inertia moment analysis of SARS CoV-2 proliferation through replication

**DOI:** 10.1101/2020.10.03.20206185

**Authors:** Vimal Raj, S Sreejyothi, M S Swapna, S Sankararaman

## Abstract

The present work proposes a surrogate method for understanding and analyzing the replication of SARS CoV-2 through fractal and inertia moment (IM) analysis of cell culture images at different stages. The fractal analysis of images of cell culture, calculated by the box-counting and power spectral density methods, reflect the stages of virus infection, leading to the replication of the virus RNA and damaging the host cell. The linear increase of IM value reveals not only the proliferation of SARS CoV-2 by replication but also damage to the host cell with time. Thus, the work shows the possibility of fractal analysis and IM measurement for understanding the dynamics of the virus infection.

## 1. Introduction

Since the dawn of 2020, the world witnessed the outbreak of Coronavirus Disease 2019 (COVID 19), a pandemic as adjudged by the World Health Organization (WHO), killing lakhs. The world struggle hard to tackle the panic situation, which is evident from the recent research publications in all realms of knowledge, crossing the barriers of science and social science [1,2]. In the current scenario, every spark of novel ideas can kindle the minds of thousands in achieving the goal. Through this paper, we propose a surrogate method for understanding and analyzing the replication of Severe Acute Respiratory Syndrome coronavirus - SARS CoV-2, employing the principles of fractal analysis and inertia moment (IM) estimation. From the exploration of images through IM and fractal signature, displayed by an object or process, its evolution kinetics can be studied [3–6]. Today, image analysis has emerged as a powerful tool of medical diagnosis [6–9] apart from is technological applications [10]. This paper is the first report of employing fractal dimension and IM values to unveil the replication kinetics of coronavirus (CoV).

The gene mappings of CoV reveal that it belongs to the Nidovirus family and betacoronavirus genus. Among the four structural proteins of CoV (Fig. 1 [11]), the Spike (S) protein attaches to the host cell and promotes the fusion between cell and virus membrane [12]. After that, it releases the virion RNA to the host cell [13] and replicates in the cytoplasm [14], leading to the destruction of the host cell and the production of new viruses. The investigation of the replication kinetics of CoVs and cell death can give valuable information in getting control over its spreading. Nowadays, image processing techniques are widely used in analyzing medical and biological problems [15]. Image analysis of cell culture is a potential tool for understanding the distribution of the cells, the kinetics of drug action, the interaction between different entities, and the quantification of specific cells [16,17]. The digital images of the cell culture are intricate patterns, whose complexity can be quantified through fractal analysis [18,19]. The analysis of the images of different stages in the cell culture, by measuring the Inertia moment (IM) can provide information about the rate of drug action and the time at which the reaction ceases [20]. IM is the second-order moment of the co-occurrence matrix that reflects even the minute changes in the images. The present study focuses on the image analysis of the kinetics of the attack of the CoV with normal cells in cell culture.

**Fig. 1.**
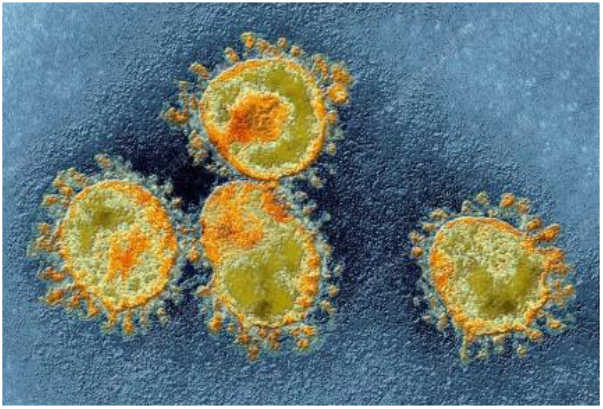
Electron micrograph of SARS CoV-2 [11].

## 2. Materials and methods

A better knowledge of the mechanism of action of the CoV and the interaction of different vaccines with it can be had by investigating the virus in a cultured medium. The scientists from the Doherty Institute in Melbourne have successfully extracted the SARS CoV-2 from a patient sample, and a video of the interaction of the virus with normal cell culture is published on their website (https://www.doherty.edu.au/news-events/news/coronavirus) [21]. In the present work, 388 images of virus interaction are obtained by splitting the video into different frames using video processing software. The images are sequentially subjected to fractal analysis and a modified IM measurement technique to study the virus interaction.

Quantification of the complexity of intricate biological structures such as cells, viruses, tissues, and others can be analyzed through fractal analysis by estimating the fractal dimension. The power spectrum, epsilon bracket, walking divider, correlation, prism counting, box-counting, and other methods are available for finding the fractal dimension of a structure. For thresholded two dimensional images, the box-counting method is a simple and effective one[5,22]. In the box-counting method, one counts the number of boxes (N(ε)), with size ‘ε’, required to fill the thresholded two-dimensional image for the different box sizes. The number of boxes and box size are related through the fractal power law,

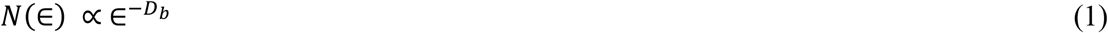

where the exponent D_b_ is the box counting fractal dimension of the structure under study. Taking the logarithm of Eq. (1) on both sides we get,

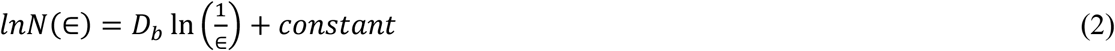

Equation (2) resembles a straight line equation, and therefore the box-counting fractal dimension ‘D_b_’ can be estimated by finding the slope of the *lnN*(∈) versus 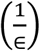 plot.

Power spectral density technique is another tool for assessing the complexity of an image from the power spectral fractal dimension (D_p_). The power spectral density technique does not require any thresholding process for the determination of D_p_. For finding the D_p_ of an image, the mean value of pixel intensities along a column is determined by sectioning it into ‘m’ rows and ‘n’ columns and then transferring it into the frequency domain using Fast Fourier Transform (FFT) technique. The square of the magnitude of the FFT signal gives the power spectral density (P). The slope (β) of ln (P) vs ln (f) gives the value of power spectral fractal dimension (D_p_) through the Eq. (3) [23,24]. Similar to the complexity mapping studies of intricate structures by finding the fractal dimensions, the study of temporal changes in these structures is also essential.

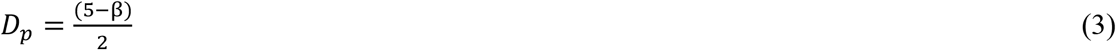

The study of the temporal evolution of a dynamical process using the IM gives significant insight into its mechanism. For calculating the IM, a matrix is made by combining the grayscale values of the central column of each frame of a dynamical process in sequential order. Then a co-occurrence matrix (M), which gives the number of occurrence (M_i,j_) of the grayscale value ‘i’ immediately followed by the grayscale value ‘j’ while moving horizontally along the new matrix, is made. The spread of the co-occurrence matrix from the diagonal is a measure of the changes in the dynamical process. From the co-occurrence matrix, the IM is calculated using the Eq. (4). The step of normalization of the co-occurrence matrix is neglected in the present study to understand the cumulative effect of the interaction of CoV with normal cells [20].

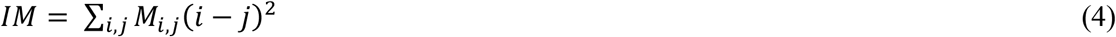

## 3. Results and discussion

The fractal dimension provides insights into the complexity of intricate cell structures. For finding the box-counting fractal dimension (D_b_), the 388 images are threshold at a level of 0.25 and then subjected to the box-counting method in Matlab software. A representative ln(N(ε)) versus ln(1/ ε) plot for finding the value of D_b_ of the first frame is shown in Fig. 2. The original image of the cell culture and its threshold image at different frames are shown in Fig. 3. The value of D_b_ of the frames is shown in Fig. 4. From Fig. 4, it can be seen that the CoV attack on cells is initiated, which is evident from the increasing complexity of the image. From the fractal analysis by box-counting method, it can be seen that the value of D_b_ increases from frame number 1 (Fig. 3a) to 32 (Fig. 3b). From literature, it can be seen that the increase in complexity gets reflected as the increasing value of the fractal dimension [18]. During the initial stage of CoV attack, the pH of the cytoplasm gets lowered. Then the CoV binds with the host receptor and fuses into the cell cytoplasm resulting in the deformation of the host cell [25]. This gets reflected in the image of cell culture as an increase in the value of D_b_. As we move from frame number 32 (Fig. 3b) to 153 (Fig. 3c), it is found that the value of D_b_ decreases indicating a decrease in complexity with respect to the frame number 32. During this period, the virus enters the host cell, gets its RNA replicated and starts killing the host cell. This replication period reflects as a decrease in the value of D_b_ from frame number 32 to 153. After that, the value of D_b_ increases from frame 153 (Fig. 3c) to frame 388 (Fig. 3d), during which the viral attack increases rapidly leading to the sharp increase in the death of host cells. Thus the fractal analysis helps in understanding the different phases of the CoV attack into a host cell. Since there is a chance of information loss during the thresholding of the image for finding the box-counting dimension, it is better to verify the results obtained by analyzing the image itself by finding its power spectral fractal dimension (D_p_). The power spectrum (ln P vs ln f) of the first frame is shown in Fig. 5a as a representative, the slope of which gives the value of D_p_ from Eq. (3). The power spectral density analysis of the 388 images gives the variation of the value of D_p_ as described in Fig. 5b. Figure 4 and 5b are complementary to each other since the former is from the thresholded images. Thus, Fig. 5b agrees well with Fig. 4 and the inference drawn.

**Fig. 2.**
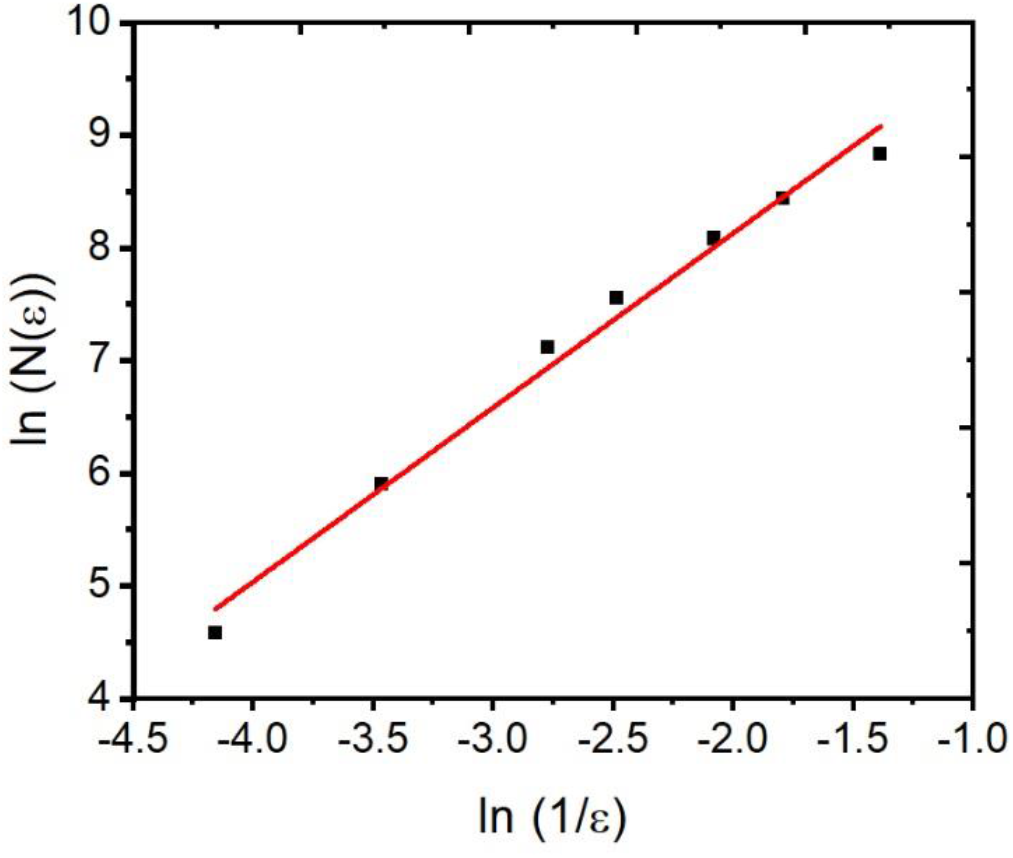
ln (N(ε)) vs ln (1/ ε) plot of the first frame.

**Fig. 3.**
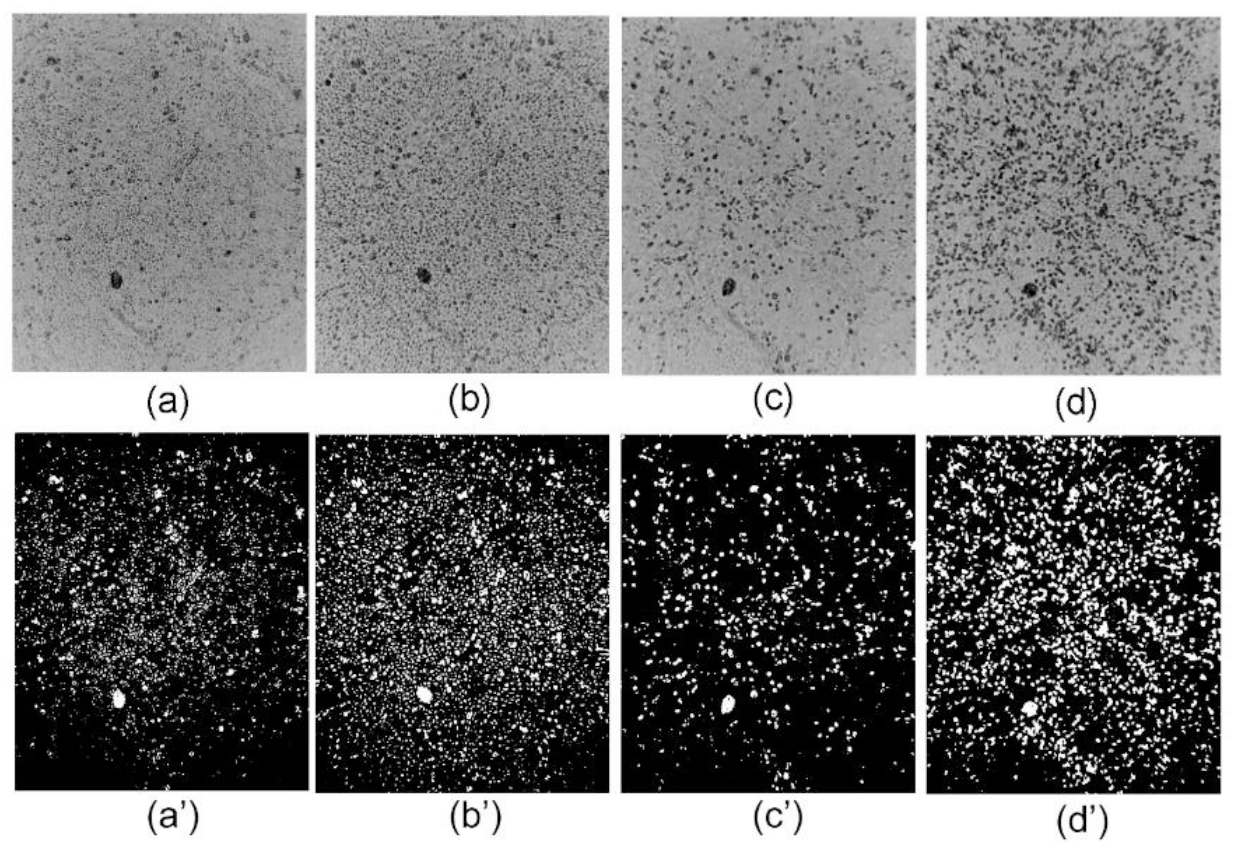
Images of the frames (a) 1, (b) 32, (c) 153, and (d) 388 with the respective threshold images (a’), (b’), (c’) and (d’).

**Fig. 4.**
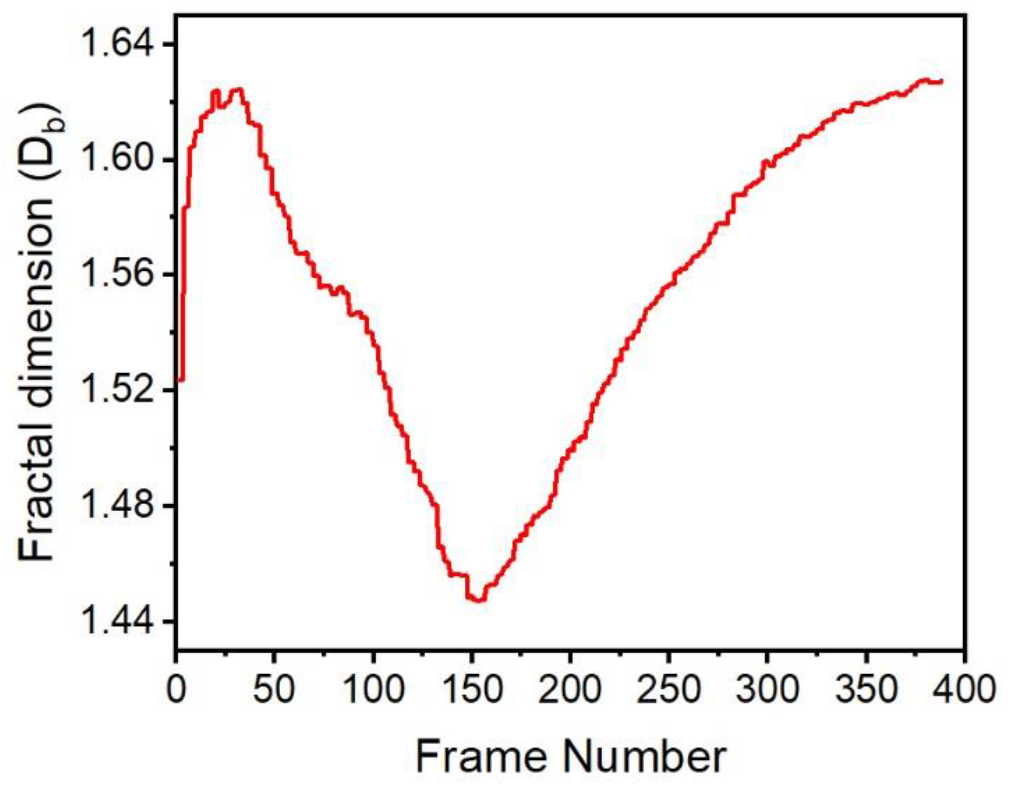
Variation of box counting fractal dimension with frame number.

**Fig. 5.**
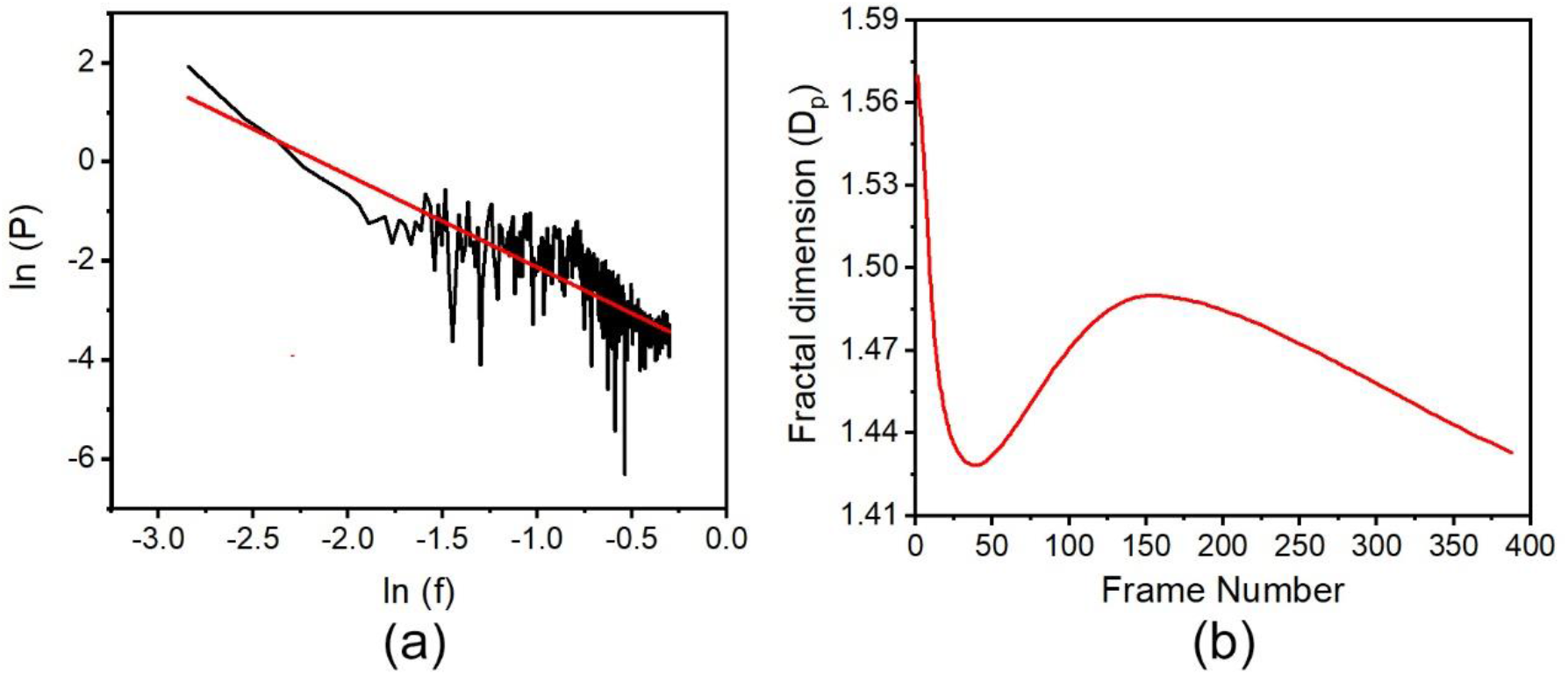
(a) ln (P) vs ln (f) plot of the first frame and (b) Variation power spectral fractal dimension with frame number.

The kinetics of a dynamical process can be understood by analyzing the co-occurrence matrix and from the value of IM. The plot of the co-occurrence matrix for frame numbers 40, 200, and 388 is shown in Fig. 6. Figure 6 shows that with the increase in time or frame number, there is a spread in the co-occurrence matrix from its diagonal. This spread implies the increased virus activity with time. The IM for different frame numbers calculated using Matlab software by taking the first frame as the reference is shown in Fig. 7. From Fig. 7, it can be seen that the IM value is increasing almost linearly with the frame number. One of the reasons for the linear increase is the fast replication of the CoV. Another reason is the increase in the dead host cell by the infection of CoV. The study shows the potential of the co-occurrence matrix plot and IM values in determining the rate of virus replication and virus infection.

**Fig. 6.**
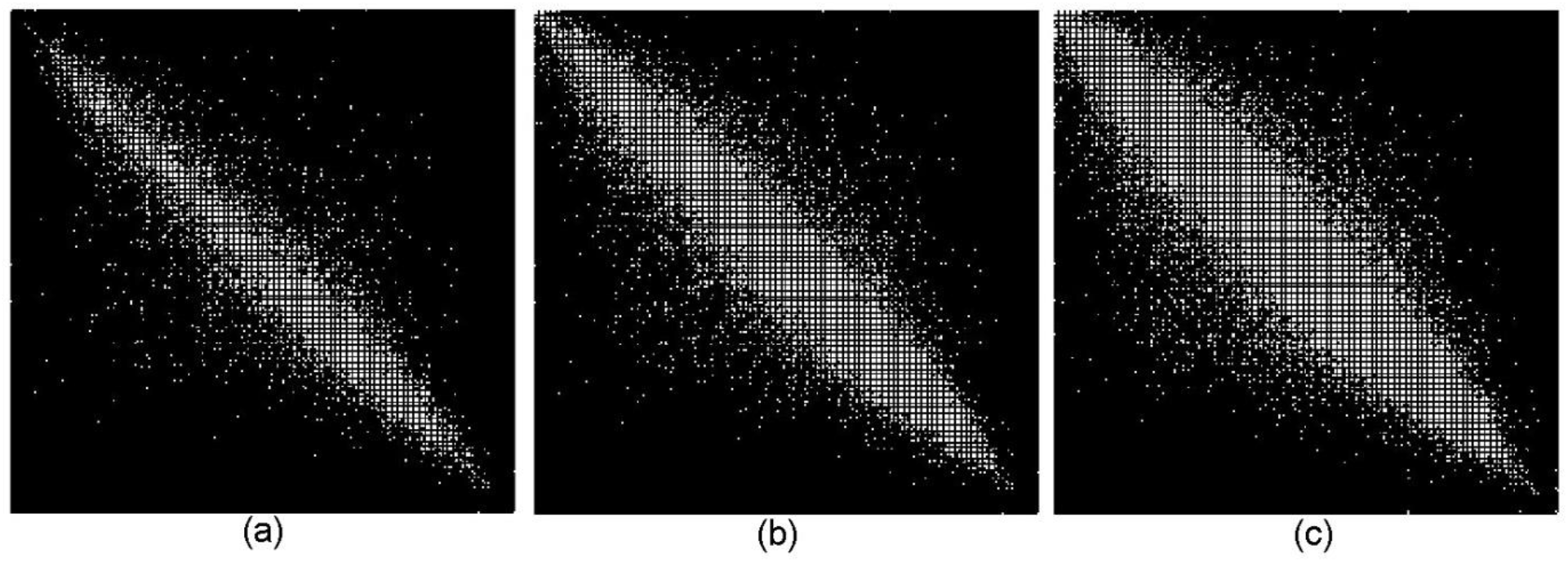
Co-occurrence matrix for the frame number (a) 40, (b) 200, and (c) 388.

**Fig. 7.**
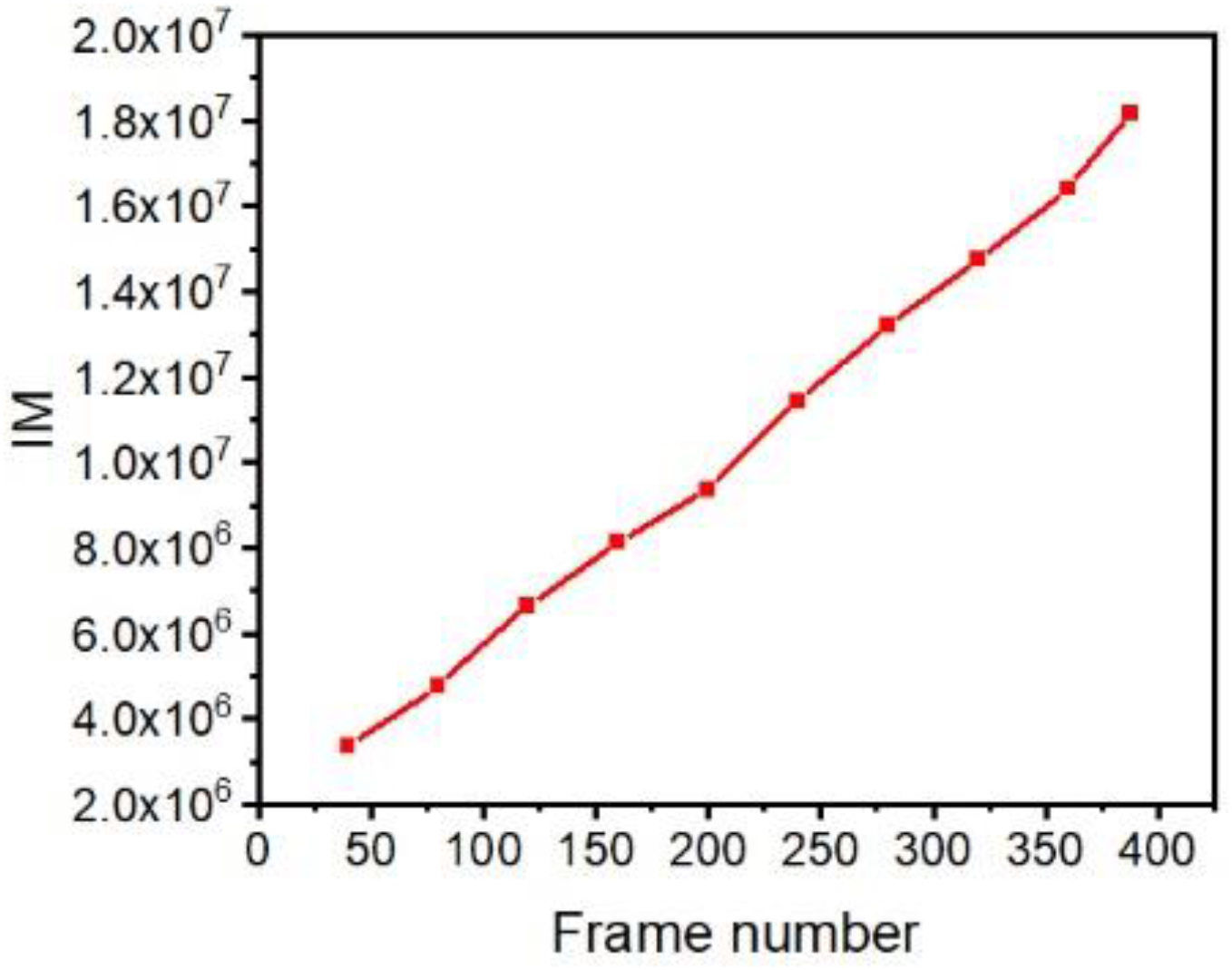
Variation of the IM with the frame number.

## 4. Conclusion

The present work proposes a surrogate method for understanding and analyzing the replication of CoV by employing the principles of fractal analysis and inertia moment (IM) estimation. A better knowledge of the mechanism of action of the CoV and the interaction of different vaccines with it can be had by investigating the virus in a cultured medium. For this, the video of the interaction of CoV with normal cell culture, published by Doherty Institute in Melbourne on their website https://www.doherty.edu.au/news-events/news/coronavirus, is subjected to fractal and IM analysis by extracting 388 image frames. The box-counting fractal dimension (D_b_) of these frames, calculated from the thresholded images, reflects the stages of virus infection resulting in the proliferation by the replication of the virus RNA and thereby damaging the living cells. The power spectral analysis of these images by finding the fractal dimension D_p_ also supports the box-counting fractal analysis. The Inertial moment value increases linearly with frame number indicating the proliferation of SARS CoV-2 through replication. Thus, the work shows the possibility of fractal analysis and IM measurement for understanding the dynamics of the virus infection.

## Data Availability

The data that support the findings of this study are available from the corresponding author upon reasonable request.

